# Predictive approaches to heterogeneous treatment effects: a systematic review

**DOI:** 10.1101/19010827

**Authors:** Alexandros Rekkas, Jessica K. Paulus, Gowri Raman, John B. Wong, Ewout W. Steyerberg, Peter R. Rijnbeek, David M. Kent, David van Klaveren

## Abstract

**Background:** Recent evidence suggests that there is often substantial variation in the benefits and harms across a trial population. We aimed to identify regression modeling approaches that assess heterogeneity of treatment effect within a randomized clinical trial.

**Methods:** We performed a literature review using a broad search strategy, complemented by suggestions of a technical expert panel.

**Results:** The approaches are classified into 3 categories: 1) Risk-based methods (11 papers) use only prognostic factors to define patient subgroups, relying on the mathematical dependency of the absolute risk difference on baseline risk; 2) Treatment effect modeling methods (9 papers) use both prognostic factors and treatment effect modifiers to explore characteristics that interact with the effects of therapy on a relative scale. These methods couple data-driven subgroup identification with approaches to prevent overfitting, such as penalization or use of separate data sets for subgroup identification and effect estimation. 3) Optimal treatment regime methods (12 papers) focus primarily on treatment effect modifiers to classify the trial population into those who benefit from treatment and those who do not. Finally, we also identified papers which describe model evaluation methods (4 papers).

**Conclusion:** Three classes of approaches were identified to assess heterogeneity of treatment effect. Methodological research, including both simulations and empirical evaluations, is required to compare the available methods in different settings and to derive well-informed guidance for their application in RCT analysis.

**Key messages:**

- Heterogeneity of treatment effect refers to the non-random variation in the direction or magnitude of a treatment effect for individuals within a population.
- A large number of regression-based predictive approaches to the analysis of treatment effect heterogeneity exists, which can be divided into three broad classes based on if they incorporate: prognostic factors (risk-based methods); treatment effect modifiers (optimal treatment regime methods); or both (treatment effect modeling methods).
- Simulations and empirical evaluations are required to compare the available methods in different settings and to derive well-informed guidance for their application in RCT analysis.

## Introduction

Evidence based medicine (EBM) has heavily influenced the standards of current medical practice. Randomized clinical trials (RCTs) and meta-analyses of RCTs are regarded as the gold standards for evidence generation within the EBM framework. Within this framework, if a patient being considered for treatment meets trial enrollment criteria, then it has been assumed that the average benefits and the benefit-harm tradeoffs are likely to apply to that individual (1). Using this reasoning, it has been argued that RCTs should attempt to include even broader populations to ensure generalizability of their results to more (and more diverse) individuals (2, 3).

However, generalizability of an RCT result and applicability to a specific patient move in opposite directions (4). When trial enrollees differ from one another in many key determinants of the outcome of interest—and consequently in the potential benefits and harms of therapy—it can be unclear to whom the overall average benefit-harm trade-offs actually apply—even among those included in the trial (5, 6). Precision medicine aims to tailor treatment to individual patients. As such, analysis of heterogeneity of treatment effect (HTE), i.e. non-random variation in the direction or magnitude of a treatment effect for individuals within a population (7), is the cornerstone of precision medicine; its goal is to predict the optimal treatments at the individual level, accounting for an individual’s risk for harm and benefit outcomes. We use the term HTE to refer to a scale dependent property (either absolute or relative) — not a property that refers only to relative changes in effect; we will specify the scale as needed. We use the term “effect modifier” and “effect modification” when we are specifically referring to variables that modify effects on a relative scale (e.g. hazard ratio or odds ratio) (8).

In this review, we focus on regression-based *predictive* approaches to HTE analysis. Such approaches predict which individual patients benefit from interventions, using all the available relevant information for that patient (9). We distinguish these analyses from the typical one-variable-at-a-time subgroup analyses that appear in forest plots of most major trial reports, and from other HTE analyses which explore or confirm causal hypotheses regarding whether a specific covariate or biomarker modifies the effects of therapy. To guide future work on individualizing treatment decisions, we aimed to summarize the methodological literature on regression modeling approaches to predictive HTE analysis.

## Methods

Due to the absence of medical subject headings for HTE, we used a relatively broad search strategy to maximize sensitivity. For the time period 1/1/2000 through 8/9/2018, we searched Medline and Cochrane Central using the text word search strategy from Table 1. We also retrieved seminal articles suggested by a technical expert panel (TEP). We reviewed the references of full-text articles meeting our eligibility criteria and retrieved full texts of citations potentially meeting our eligibility criteria.

**Table 1:** Search strategy for the study.

| # | Results |
| --- | --- |
| 1 | ((heterogen\$ and effect\$) or (effect and modif\$)).tw. |
| 2 | regression.tw. |
| 3 | treatment\$.tw. |
| 4 | (treatment adj1 effect\$).tw. |
| 5 | (treatment adj1 difference\$).tw. |
| 6 | exp risk/ or risk.tw. |
| 7 | 3 or 4 or 5 or 6 |
| 8 | *Models, Statistical/ |
| 9 | *Randomized Controlled Trials as Topic/mt |
| 10 | Multicenter Studies as Topic/mt |
| 11 | *Randomized Controlled Trials as Topic/sn |
| 12 | Multicenter Studies as Topic/sn |
| 13 | *Clinical Trials as Topic/sn |
| 14 | *Precision Medicine/mt |
| 15 | or/8-14 |
| 16 | 1 and 2 |
| 17 | 2 and 7 |
| 18 | 15 and 17 |
| 19 | 15 and 16 |
| 20 | 18 or 19 |

We sought papers that developed or evaluated methods for predictive HTE in the setting of parallel arm RCT designs or simulated RCT. Abstracts were screened to identify papers that developed or evaluated a regression-based method for predictive HTE on actual or simulated parallel arm RCT data. Papers describing a generic approach that could be applied using either regression or non-regression methods, or papers comparing regression to non-regression methods were also included. Similarly, papers comparing generic one-variable-at-a-time approaches to predictive HTE methods were also included. Finally, papers suggested by the TEP that fell outside the search window were considered for inclusion.

We excluded papers solely related to cross-over, single-arm, and observational study designs. Papers applying predictive HTE methods only to address clinical aims were excluded. We also rejected papers using only non-regression-based methods. Similarly, methods papers about ONLY non-predictive subgroup analysis, i.e. one-variable-at-a-time or conventional subgroups, were omitted. We excluded papers on trial enrichment or adaptive trial designs along with those that use predictive HTE approaches in the design. We also excluded papers primarily aiming at characterization or identification of heterogeneity in response rather than trying to predict responses for individual patients or subsets of patients; e.g. group based trajectory or growth mixture modeling. Papers on regression methods that make use of covariates post-baseline, or temporally downstream of the treatment decision were omitted. Review articles and primarily conceptual papers without accompanying methods development were also excluded.

Titles and abstracts were retrieved and double-screened by six independent reviewers against eligibility criteria. Disagreements were resolved by group consensus in consultation with a seventh senior expert reviewer (DMK) in meetings.

## Results

We identified 2510 abstracts that were screened in duplicate. We retrieved 64 full-text articles and an additional 110 suggested by experts and identified from reference lists of eligible articles. These 174 full-text articles were again screened in duplicate with group consensus resolution of conflicts in meetings. A total of 36 articles met eligibility criteria (Figure 1).

**Figure 1:**
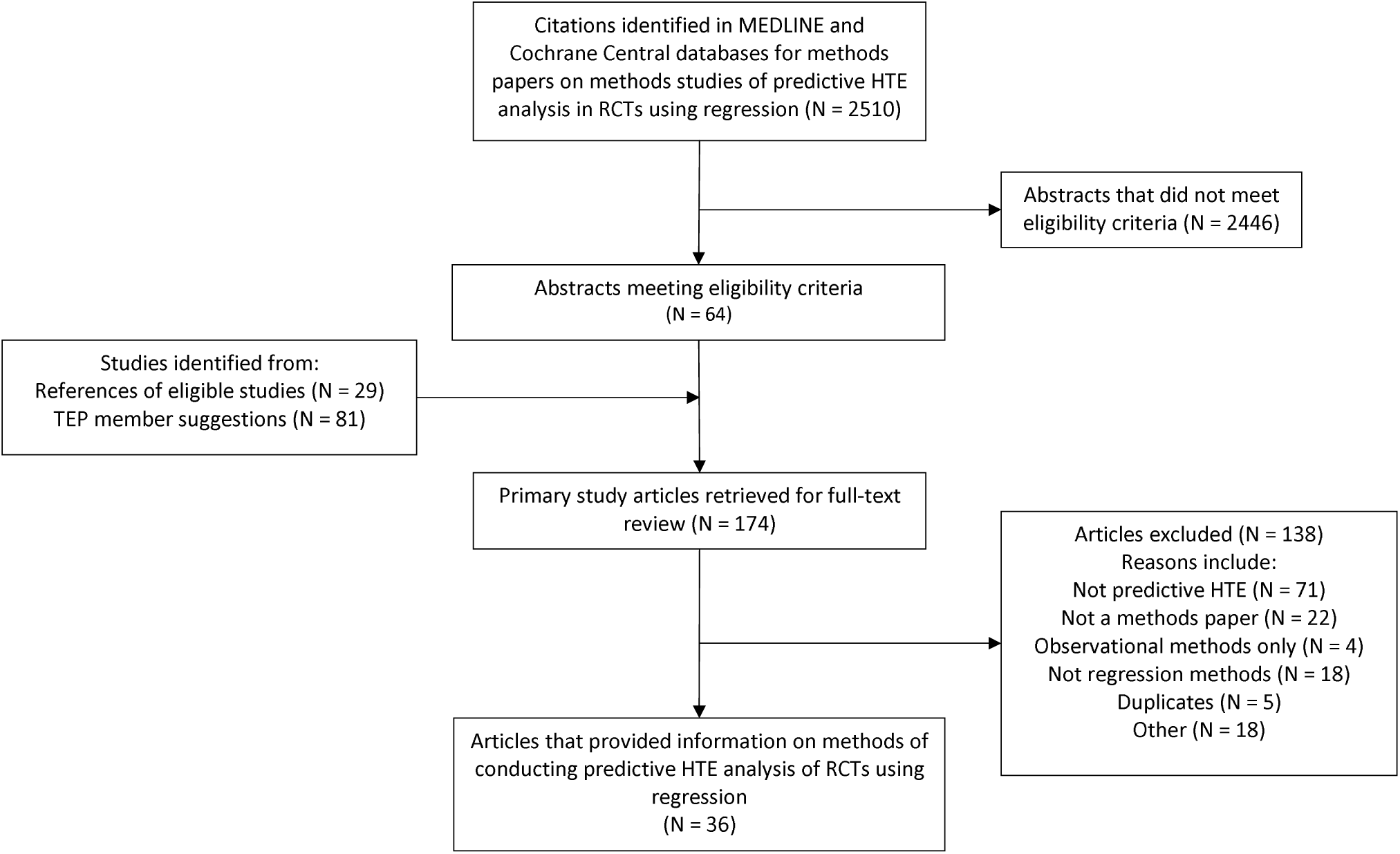
Study flow chart.

### Categorization methods

We could classify all regression-based methods to predictive HTE into 3 broad categories based on whether and how they incorporated prognostic variables and relative treatment effect modifiers:

- **Risk-based methods** exploit the mathematical dependency of treatment benefit on a patient’s baseline risk. Even though relative treatment effect may vary across different levels of baseline risk, relative treatment effect modification by each covariate is not considered (Table 2, equations 1-3).
- **Treatment effect modeling methods** use both the main effects of risk factors and covariate-by-treatment interaction terms (on the relative scale) to estimate individualized benefits. They can be used either for making individualized absolute benefit predictions or for defining patient subgroups with similar expected treatment benefits (Table 2, equation 4).
- **Optimal treatment regime methods** focus primarily on treatment effect modifiers for the definition of a treatment assignment rule dividing the trial population into those who benefit from treatment and those who do not (Table 2, equation 5). Contrary to previous methods, baseline risk or the magnitude of absolute treatment benefit are not of primary concern.

**Table 2:**
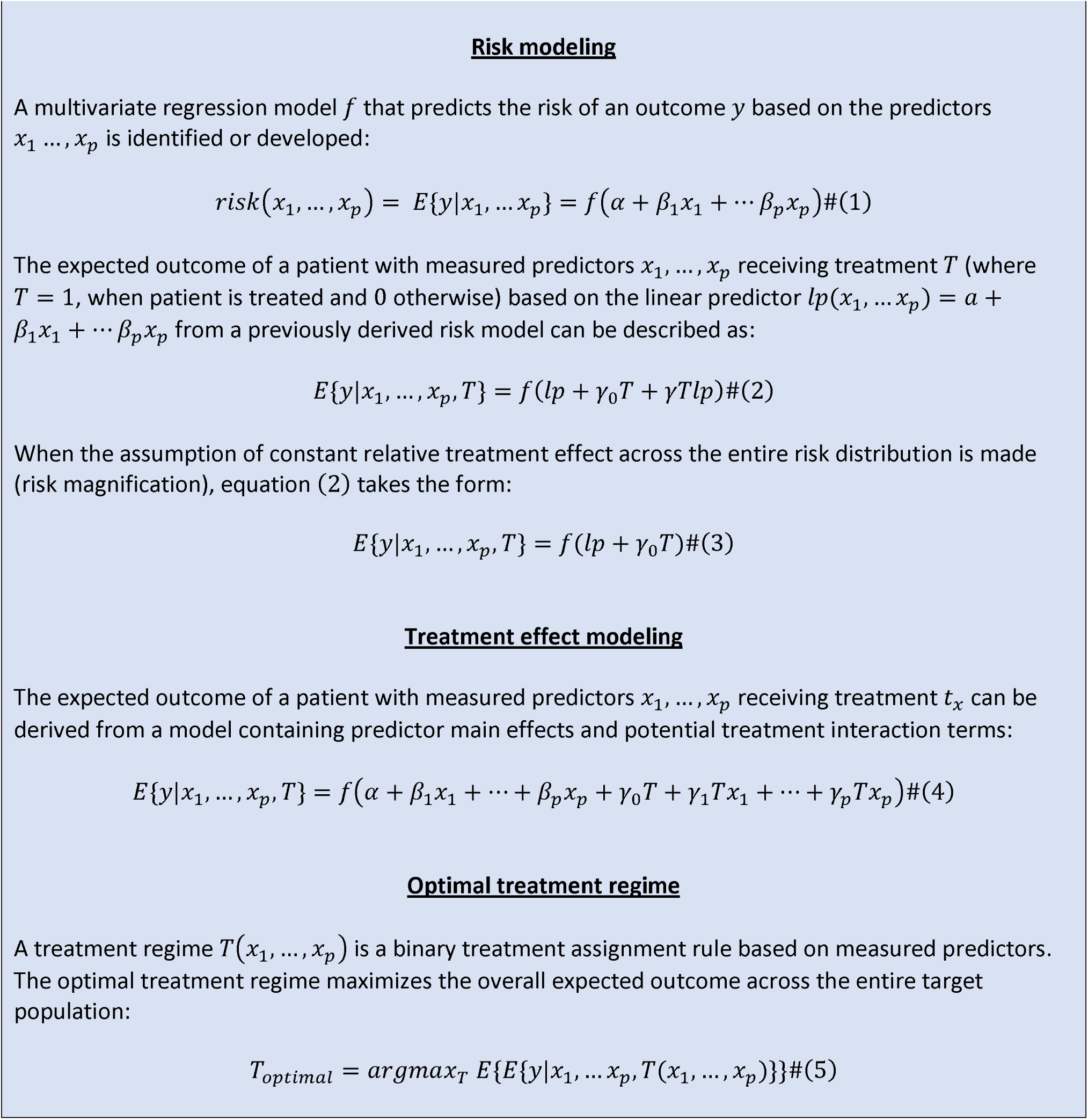
Equations corresponding to treatment effect heterogeneity assessment methods.

Although risk-based methods emerged earlier (Figure 2), methodology papers on treatment effect modeling (9 papers) and optimal treatment regimes (12 papers) are more frequently published since 2010 than papers on risk-based methods (8 papers). Even though extensive literature exists on model evaluation when it comes to prediction modeling, the same task can be quite challenging when modeling treatment effects (10). That is due to the unavailability of counterfactual outcomes under the alternative treatment, providing a substantial challenge to the assessment of model fit. Methods included in the review concerning model evaluation in the setting of predictive HTE (4 papers) were assigned to a separate category as they are relevant to all identified approaches.

**Figure 2:**
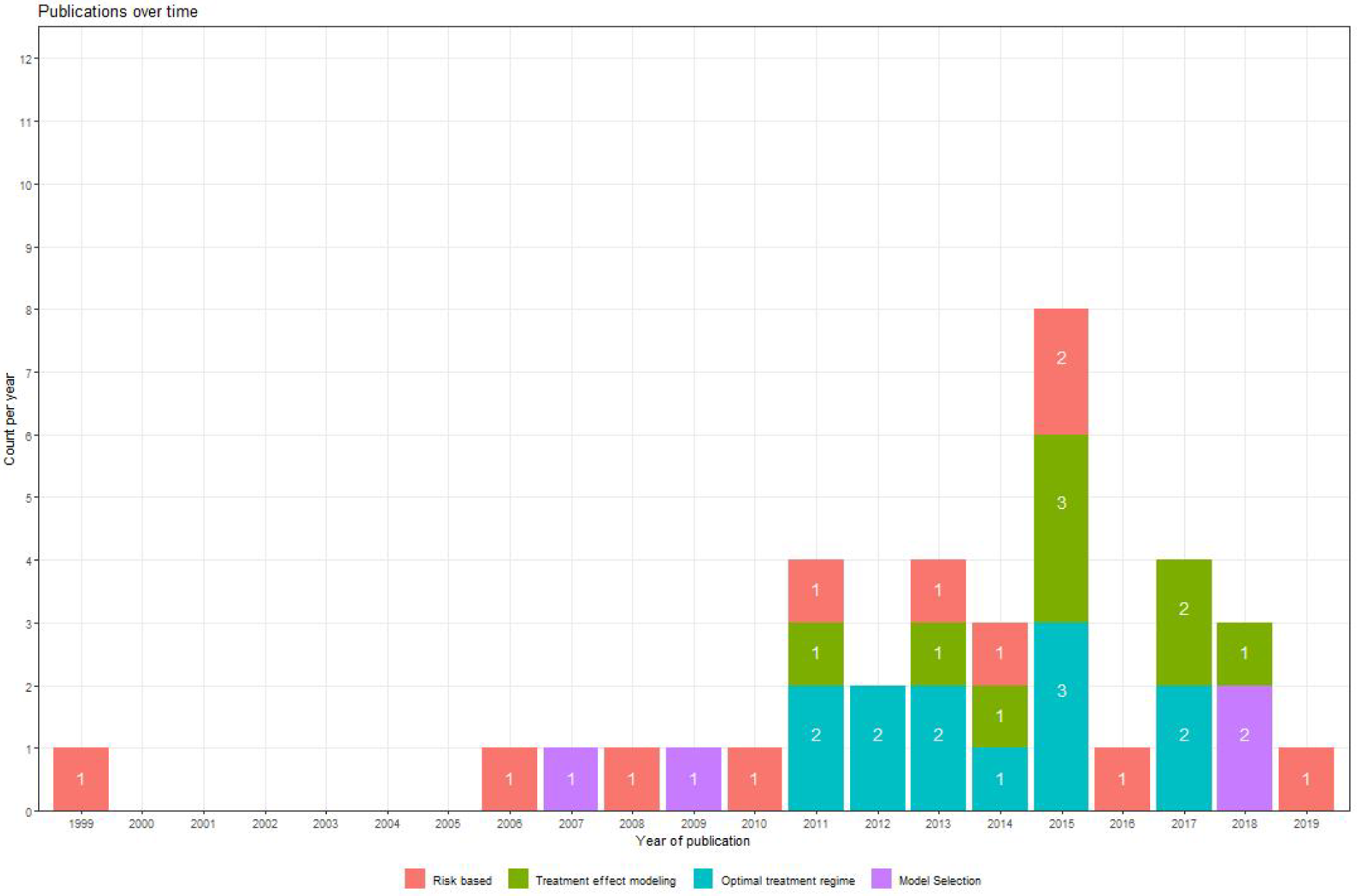
Publications over time. Publications included in the review from 1999 until 2019. Numbers inside the bars indicate the method-specific number of publications made in a specific year.

### Risk-based methods

The most rigid and straightforward risk-based methods assume a constant relative treatment effect across different levels of baseline risk and ignore potential interactions with treatment. Dorresteijn et al. (11) studied individualized treatment with rosuvastatin for prevention of cardiovascular events. They combined existing prediction models (Framingham score, Reynolds risk score) with the average rosuvastatin effect found in an RCT. To obtain individualized absolute treatment benefits, they multiplied baseline risk predictions with the average risk reduction found in trials. The value of the proposed approach is assessed in terms of improved decision making by comparing the net benefit with treat-none and treat-all strategies (12). Julien and Hanley (13) estimated prognostic effects and treatment effect directly form trial data, by incorporating a constant relative treatment effect term in a Cox regression model. Patient-specific benefit predictions followed from the difference between event-free survival predictions for patients with and without treatment. A similar approach was used to obtain the predicted 30-day survival benefit of treatment with aggressive thrombolysis after acute myocardial infarction (14).

Risk stratification approaches analyze relative treatment effects and absolute treatment effects within strata of predicted risk, rather than assuming a constant relative effect. Both Hayward et al. (15) and Iwashyna et al. (16) demonstrated that these methods are useful in the presence of treatment-related harms to identify patients who do not benefit (or receive net harm) from a treatment that is beneficial on average. In a range of plausible scenarios, simulations showed that studies were generally underpowered to detect covariate-by-treatment interactions, but adequately powered to detect risk-by-treatment interactions, even when a moderately performing prediction model was used to stratify patients. Hence, risk stratification methods can detect patient subgroups that have net harm even when conventional methods conclude consistency of effects across all major subgroups.

Kent et al. (17) proposed a framework for HTE analysis in RCT data that recommended published trials routinely report the distribution of baseline risk in the overall study population and in the separate treatment arms using a risk prediction tool. Researchers should demonstrate how relative and absolute risk reduction vary by baseline risk and test for HTE with interaction tests. Externally validated prediction models should be used, when available.

In the absence of an adequate prediction model when performing a risk-based assessment of HTE, an internal risk model from the data at hand can be derived. Burke et al. (18) demonstrated that developing the risk model on the control arm of the trial may result in overfitting and suboptimal risk-based assessment of HTE, exaggerating its presence. In extensive simulations, internally developed prediction models blinded to treatment assignment led to unbiased treatment effect estimates in strata of predicted risk. Using this approach to re-analyze 32 large RCT, Kent et al. (19) demonstrated that there is virtually always substantial variation in outcome risk within an RCT, which in turn leads to substantial HTE on the clinically important scale of absolute risk difference. Several trials from this analysis had clinically relevant results (20-22).

Similar to Burke et al. (18), Abadie et al. (23) presented evidence of large biases in risk stratified assessment of HTE in two randomized experiments rising from the development of a prediction model solely from the control arm. As a remedy, they considered both a leave-one-out approach, where individual risk predictions are obtained from a model derived by excluding the particular individual, and a repeated split sample approach, where the original sample is repeatedly split into a sample for the development of the prediction model and a sample for treatment effect estimation within risk strata. These approaches were found to substantially reduce bias in a simulation study. Finally, Groenwold et al. (24) found in simulations that the inclusion of a constant relative treatment effect in the development of a prediction model better calibrates predictions to the untreated population. However, this approach may not be optimal for risk-based assessment of HTE, where accurate ranking of risk predictions is of primary importance for the calibration of treatment benefit predictions.

Follmann and Proschan (25) proposed a one-step likelihood ratio test procedure based on a proportional interactions model to decide whether treatment interacts with a linear combination of baseline covariates. Their proportional interactions model assumes that the effects of prognostic factors in the treatment arm are equal to their effects in the control arm multiplied by a constant, the proportionality factor. Testing for an interaction along the linear predictor amounts to testing that the proportionality factor is equal to 1. If high risk patients benefit more from treatment (on the relative scale) and disease severity is determined by a variety of prognostic factors, the proposed test results in greater power to detect HTE on the relative scale compared to multiplicity-corrected subgroup analyses.

Kovalchik et al. (26) expanded upon the previous approach by exploring misspecification of the proportional interactions model, when considering a fixed set of pre-specified candidate effect modifiers. A proportional interactions model is misspecified either when covariates with truly proportional effects are excluded or when covariates with non-proportional effects across treatment arms are included in the model. In this case the one-step likelihood ratio test of Follmann and Proschan (25) fails to achieve its statistical advantages. For model selection an all subsets approach combined with a modified Bonferroni correction method can be used. This approach accounts for correlation among nested subsets of considered proportional interactions models, thus allowing the assessment of all possible proportional interactions models while controlling for the family-wise error rate.

### Treatment effect modeling

Using data from the SYNTAX trial (27) Van Klaveren et al. (28) considered models of increasing complexity for the assessment of HTE at the individual level using data from the SYNTAX trial. They compared different Cox regression models for the prediction of treatment benefit: 1) a model without any risk factors; 2) a model with risk factors and a constant relative treatment effect; 3) a model with treatment, a prognostic index and their interaction; and 4) a model including treatment interactions with all available prognostic factors, fitted both with conventional and with penalized ridge regression. Benefit predictions at the individual level were highly dependent on the modeling strategy, with treatment interactions improving treatment recommendations under certain circumstances.

Basu et al. (29) developed and validated risk models for predicting the absolute benefit (reduction of CVD events) and harm (serious adverse events) from intensive blood pressure therapy, using data from SPRINT. They compared traditional backward selection to an elastic net approach for selection and estimation of all treatment-covariate interactions. The two approaches selected different treatment-covariate interactions and—while their performance in terms of CVD risk prediction was comparable when externally validated in the ACCORD BP trial (30)—the traditional approach performed considerably worse than the penalized approach when predicting absolute treatment benefit. However, with regard to selection of treatment interactions, Ternes et al. (31) concluded from an extensive simulation study that no single methodology yielded uniformly superior performance. They compared 12 different approaches in a high-dimensional setting with survival outcomes. Their methods ranged from a straightforward univariate approach as a baseline, where Wald tests accounting for multiple testing were performed for each treatment-covariate interaction to different approaches for dealing with hierarchy of effects—whether they enforce the inclusion of the respective main effects if an interaction is selected—and also different magnitude of penalization of main and interaction effects.

Another approach to reducing overfitting of treatment effect models is separation of treatment effect estimation from subgroup identification. Cai et al. (32) fit “working” regression parametric models within treatment arms to derive absolute treatment benefit scores initially. In a second stage, the population is stratified into small groups with similar predicted benefits based on the first-stage scores. A non-parametric local likelihood approach is used to provide a smooth estimate of absolute treatment benefit across the range of the derived sores. Claggett et al. (33) extended this two-stage methodology to RCTs with multiple outcomes, by assigning outcomes into meaningful ordinal categories. Overfitting can be avoided by randomly splitting the sample into two parts; the first part is used to select and fit ordinal regression models in both the treatment and the control arm. In the second part, the models that perform best in terms of a cross-validated estimate of concordance between predicted and unobservable true treatment difference— defined as the difference in probability of observing a worse outcome under control compared to treatment and the probability of observing a worse outcome under treatment compared to control—are used to define treatment benefit scores for patients. Treatment effects conditional on the treatment benefit score are then estimated through a non-parametric kernel estimation procedure.

Zhao et al. (34) proposed a two-stage methodology similar to the approach of Cai et al. (32), focusing on the identification of a subgroup that benefits from treatment. They repeatedly split the sample population based on the first-stage treatment benefit scores and estimate the treatment effect in subgroups above different thresholds. These estimates are plotted against the score thresholds to assess the adequacy of the selected scoring rule. This method could also be used for the evaluation of different modeling strategies by selecting the one that identifies the largest subgroup with an effect estimate above a desired threshold.

Künzel et al. (35) proposed an “X-learner” for settings where one treatment arm is substantially larger than the alternative. They also start by fitting separate outcome models within treatment arms. However, rather than using these models to calculate treatment benefit scores, they imputed individualized absolute treatment effects, defined as the difference between the observed outcomes and the expected counterfactual (potential) outcomes based on model predictions. In a second stage, two separate regression models—one in each treatment arm—are fitted to the imputed treatment effects. Finally, they combined these two regression models for a particular covariate pattern by taking a weighted average of the expected treatment effects.

Most effect modeling methods start with outcome predictions conditional on treatment and then examine the difference in predictions with and without treatment. In contrast, Weisberg and Pontes (36) introduced a causal difference outcome variable (“cadit”) which can be modeled directly. In case of a binary outcome, the binary cadit is 1 when a treated patient has a good outcome or when an untreated patient does not, and 0 otherwise. Thus, the dependent variable implicitly codes treatment assignment and outcome simultaneously. They first demonstrated that the absolute treatment benefit equals 2x P(cadit = 1) − 1 and then they derived patient-specific treatment effect estimates by fitting a logistic regression model to the cadit. A similar approach was described for continuous outcomes with the continuous cadit defined as −2 and 2 times the centered outcome, i.e. the outcome minus the overall average outcome, for untreated and treated patients, respectively.

Finally, Berger et al. (37) proposed a Bayesian methodology for the detection of subgroup treatment effects in case of a continuous response and binary covariates. The approach identifies single covariates likely to modify treatment effect, along with the expected individualized treatment effect. The authors also extended their methodology to include two covariates simultaneously, allowing for the assessment of multivariate subgroups.

### Optimal treatment regime methods

A treatment regime (TR) is a function mapping each patient’s covariate pattern to a single treatment assignment. Any candidate TR can be evaluated based on its value, i.e. the expected outcome at the population level if the specific TR were to be followed. The TR achieving the highest value among all possible TRs is the optimal treatment regime (OTR). The majority of such methods follows a two-stage approach, where an outcome model—usually including treatment interactions—is used to derive expected treatment benefit in the first stage. In the second stage treatment assignment is optimized based on the expected outcome. Qian and Murphy (38) advocated a first-stage model including all covariate main effects and treatment interactions in combination with LASSO-penalization to reduce model complexity.

When the outcome model is misspecified, however, the approach of Qian and Murphy may fail to identify the best possible treatment regime. As Zhang et al. (39) introduced an approach robust to such misspecifications that uses an augmented inverse probability weighted estimator of the value function. This is achieved by imposing a missing data framework, where the response under any candidate OTR is observed if the proposed treatment coincides with actual treatment and is considered missing otherwise. However, in commenting on this work, Taylor et al. (40) noted that the misspecification issues of the outcome models considered in the simulation study presented by Zhang et al. would have been easily spotted, if common approaches for the assessment of model fit had been examined. They argue that if adequately fitting outcome models had been thoroughly sought, the extra modeling required for the robust methods of Zhang et al. may not have been necessary.

Zhang et al. (41) proposed a novel framework for the derivation of OTRs, within which treatment assignment is viewed as a classification problem. The OTR is derived in two separate steps. In the first step, a contrast function is estimated, determining the difference between expected outcomes under different treatment assignments for each individual patient. The sign of the contrast function is then used to define class labels, i.e. −1 for negative contrast (harm) and +1 for positive contrast (benefit). In the second step, any classification technique can be used to find the OTR by minimizing the expected misclassification error weighted by the absolute contrast. The authors demonstrated that many of the already existing OTR methods (38, 39) fit within their framework by defining a specific contrast function.

When the outcome of interest is continuous, the magnitude of absolute treatment benefit estimates derived from regression-based methods depends solely on treatment interactions. Therefore, Foster et al. (42) focus on non-parametric estimation of the function defining the structure of treatment-covariate interactions for a continuous outcome of interest. More specifically, they recursively update non-parametric estimates of the treatment-covariate interaction function from baseline risk estimates and vice-versa until convergence. The estimates of absolute treatment benefit are then used to restrict treatment to a contiguous sub-region of the covariate space.

Xu et al. (43) claimed that the identification of an OTR with high value depends on the adequate assessment of the sign of treatment-covariate interactions rather than on the estimation of the contrast function. They demonstrated that in many common cases (binary or time-to-event outcomes), even though the underlying structure of interactions can be quite complex, its sign can be approximated from a much simpler linear function of effect modifiers. Using the classification framework of Zhang et al. (41), they assign patients to class labels based on the resulting sign from these candidate linear combinations. The coefficients of that linear function are derived by minimizing the misclassification error weighted by the observed outcome—assuming higher values are preferable. In this way, the derived OTR is forced to contradict actual treatment assignment when the observed outcome is low. Tian et al. (44) proposed a different approach that solely focuses on treatment-covariate interactions by recoding the binary treatment indicator variable to −1/2 for control patients and +1/2 for treated patients and multiplying it with the covariates of a posited regression model to derive modified covariates so that the linear predictor of the model predicting the outcome from the modified covariates can be used as a score for stratifying patients with regard to treatment benefits.

Kraemer (45) suggested a methodology that implicitly assesses treatment-covariate interactions using the correlation coefficient of the pairwise difference of the outcome between treatment arms and their respective candidate predictive factor pairwise difference as a measure of effect modification. A stronger composite treatment effect modifier can then be constructed by fitting a regression model predicting pairwise outcome differences between treatments from the averages of the effect modifier values across treatment arms and then summing the individual effect modifiers weighted by the estimated regression coefficients. Treatment can then be assigned based on stratification on the composite treatment effect moderator. Two different approaches to model selection in Kraemer’s effect modifier combination method were identified in clinical applications. Principal component analysis was used to select an uncorrelated subset from a large set of possibly correlated effect modifiers (46). Alternatively, the cross-validated mean squared error of increasingly complex regression models was used to select the number of effect modifiers to construct the composite one (47).

Gunter et al. (48) proposed a method for the discovery of covariates that qualitatively interact with treatment. Using LASSO regression to reduce the space of all possible combinations of covariates and their interaction with treatment to a limited number of covariate subsets, their approach selects the optimal subset of candidate covariates by assessing the increase in the expected response from assigning based on the considered treatment effect model, versus the expected response of treating everyone with the treatment found best from the overall RCT result. The considered criterion also penalizes models for their size, providing a tradeoff between model complexity and the increase in expected response.

Finally, Petkova et al. (49) proposed to combine baseline covariates into a single generated effect modifier (GEM) based on the linear model. The GEM is defined as the linear combination of candidate effect modifiers and the objective is to derive their individual weights. This is done by fitting linear regression models within treatment arms where the independent variable is a weighted sum of the baseline covariates, while keeping the weights constant across treatment arms. The intercepts and slopes of these models along with the individual covariate GEM contributions are derived by maximizing the interaction effect in the GEM model, or by providing the best fit to the data, or by maximizing the statistical significance of an F-test for the interaction effects—a combination of the previous two. The authors derived estimates that can be calculated analytically, which makes the method easy to implement.

A growing literature exists on estimating the effect of introducing the OTR to the entire population (50-53). Luedtke and Van der Laan (50) provide an estimate of the optimal value—the value of the OTR— that is valid even when a subset of covariates exists for which treatment is neither beneficial nor harmful. It has been previously demonstrated that estimation of the optimal value is quite difficult in those situations (54). Based on the proposed method, an upper bound of what can be hoped for when a treatment rule is introduced can be established. In addition, Luedtke and Van der Laan (53) provided an estimation method for the impact of treating the optimal subgroup, i.e. the subgroup that is assigned treatment based on the OTR. Their methodology returns an estimate of the population level effect of treating based on the OTR compared to treating no one.

### Model evaluation

Schuler et al. (55) defined three broad classes of metrics relevant to model selection when it comes to treatment effect modeling. *μ*-risk metrics evaluate the ability of models to predict the outcome of interest conditional on treatment assignment. Treatment effect is either explicitly modeled by treatment interactions or implicitly by developing separate models for each treatment arm. *τ*-risk metrics focus directly on absolute treatment benefit. However, since absolute treatment benefit is unobservable, it needs to be estimated first. Value-metrics originate from OTR methods and evaluate the outcome in patients that were assigned to treatment in concordance with model recommendations.

Vickers et al. (12) suggested a methodology for the evaluation of models predicting individual treatment effects. The method relies on the expression of disease-related harms and treatment-related harms on the same scale. The minimum absolute benefit required for a patient to opt for treatment (treatment threshold) can be viewed as the ratio of treatment-related harms and harms from disease-related events, thus providing the required relationship. Net benefit is then calculated as the difference between the decrease in the proportion of disease-related events and the proportion of treated patients multiplied by the treatment threshold. The latter quantity can be viewed as harms from treatment translated to the scale of disease-related harms. Then, the net benefit of a considered prediction model at a specific treatment threshold can be derived from a patient-subset where treatment received is congruent with treatment assigned based on predicted absolute benefits and the treatment threshold. The model’s clinical relevance is derived by comparing its net benefit to the one of a treat-all policy.

Van Klaveren et al. (56) defined a measure of discrimination for treatment effect modeling. A model’s ability to discriminate between patients with higher or lower benefits is challenging, since treatment benefits are unobservable in the individual patient (since only one of two counterfactual potential outcomes can be observed). Under the assumption of uncorrelated counterfactual outcomes, conditional on model covariates, the authors matched patients from different treatment arms by their predicted treatment benefit. The difference of the observed outcomes between the matched patient pairs (0, 1: benefit; 0,0 or 1, 1: no effect; 1, 0: harm) acts as a proxy for the unobservable absolute treatment difference. The c-statistic for benefit can then be defined on the basis of this tertiary outcome as the proportion of all possible pairs of patient pairs in which the patient pair observed to have greater treatment benefit was predicted to do so.

Finally, Chen et al. (57) focused on the case when more than one outcomes—often non-continuous— are of interest and proposed a Bayesian model selection approach. Using a latent variable methodology, they link observed outcomes to unobservable quantities, allowing for their correlated nature. To perform model selection, they derive posterior probability estimates of false inclusion or false exclusion in the final model for the considered covariates. Following the definition of an outcome-space sub-region that is considered beneficial, individualized posterior probabilities of belonging to that beneficial sub-region can be derived as a by-product of the proposed methodology.

## Discussion

We identified 36 methodological papers in recent literature that describe predictive regression approaches to HTE analysis in RCT data. These methodological papers aimed to develop models for predicting individual treatment benefit and could be categorized as follows: 1) risk modeling (n=11), in which RCT patients were stratified or grouped solely on the basis of prognostic models; 2) effect modeling (n=9), in which patients are grouped or stratified by models combining prognostic factors with factors that modify treatment effects on the relative scale (effect modifiers); 3) optimal treatment regimes (n=12), which seek to classify patients into those who benefit and those who do not, primarily on the basis of effect modifiers. Papers on the evaluation of different predictive approaches to HTE (n=4) were assigned to a separate category. Of note, we also found literature on the evaluation of biomarkers for treatment selection, which did not meet inclusion criteria (58-61).

Risk-based approaches use baseline risk to define the reference class of a patient when assessing individual HTE. Two distinct approaches were identified: 1) risk magnification (62, 63) assumes constant relative treatment effect across all patient subgroups, while 2) risk stratification analyzes treatment effects within strata of predicted risk. This approach is straightforward to implement, and may provide adequate assessment of HTE in the absence of strong prior evidence for potential effect modification. The approach might better be labeled ‘benefit magnification’, since benefit increases by higher baseline risk and a constant relative risk.

Treatment effect modeling methods focus on predicting the absolute benefit of treatment through the inclusion of treatment-covariate interactions alongside the main effects of risk factors. However, modeling such interactions can result in serious overfitting of treatment benefit, especially in the absence of well-established treatment effect modifiers. Penalization methods such as LASSO regression, ridge regression or a combination (elastic net penalization) can be used as a remedy when predicting treatment benefits in other populations. Staging approaches starting from—possibly overfitted— “working” models predicting absolute treatment benefits that can later be used to calibrate predictions in groups of similar treatment benefit provide another alternative. While these approaches should yield well calibrated personalized effect estimates when data are abundant, it is yet unclear how broadly applicable these methods are in conventionally sized randomized RCTs. Similarly, the additional discrimination of benefit of these approaches compared to the less flexible risk modeling approaches remains uncertain. Simulations and empirical studies should be informative regarding these questions.

The similarity of OTRs to general classification problems—finding an optimal dichotomization of the covariates space—enables the implementation of several existing non-regression-based classification algorithms. For instance Zhao et al. (64) applied a support vector machine methodology for the derivation of an OTR for a binary outcome and was later extended to survival outcomes (65). Because prognostic factors do not affect the sign of the treatment effect, several OTR methods rely primarily on treatment effect modifiers. However, when treatments are associated with adverse events or treatment burdens (such as costs) that are not captured in the primary outcome—as is often the case—estimates of the magnitude of treatment effect are required to ensure that only patients above a certain expected net benefit threshold (i.e. outweighing the harms and burdens of therapy) are treated. Similarly, these classification methods do not provide comparable opportunity for incorporation of patient values and preferences for shared decision making which prediction methods do.

We identify several limitations to our study. Because no MeSH identifying these methods exists, we anticipate that our search approach likely missed some studies. In addition, a recently growing literature of other non-regression based methods that assess predictive HTE in observational databases (66-68) would have been excluded. Finally, our review is descriptive and did not compare the approaches for their ability to predict individualized treatment effects or to identify patient subgroups with similar expected treatment benefits

Based on the findings and the limitations of our review, several objectives for future research can be described. Optimal approaches to the reduction of overfitting through penalization need to be determined, along with optimal measures to evaluate models intended to predict treatment effect. General principles to judge the adequacy of sample sizes for predictive analytic approaches to HTE are required to complement the previous objectives. Also, methods that simultaneously predict multiple risk dimensions regarding both primary outcome risks and treatment-related harms need to be explored. The current regression-based collection of methods could be expanded by a review of non-regression approaches. Methods targeted at the observational setting need also to be considered. Additionally, a set of empirical and simulation studies should be performed to evaluate and compare the identified methods under settings representative of real world trials. The growing availability of publicly available randomized clinical trials should support this methodological research (69-71).

While there is an abundance of proposed methodological approaches, examples of clinical application of HTE prediction models remain very rare. This may reflect the fact that all these approaches confront the same fundamental challenges. These challenges include the unobservability of individual treatment response, the curse of dimensionality from the large number of covariates, the lack of knowledge about the causal molecular mechanisms underlying variation in treatment effects and the relationship of these mechanism to observable variables, and the very low power in which to explore interactions. Because of these challenges there might be very serious constraints on the usefulness of these methods as a class; while some methods may be shown to have theoretical advantages, the practical import of these theoretical advantages may not be ascertainable.

The methods we identified here generally approach the aforementioned challenges from opposite ends. Relatively rigid methods, such as risk magnification (in which relative effect homogeneity is assumed) and risk modeling (which examines changes in relative effect according to baselines risk only) deal with dimensionality, low power and low prior knowledge by restricting the flexibility of the models that can be built to emphasize the well understood influence of prognosis. Effect modeling approaches permit more flexible modeling and then subsequently try to correct for the overfitting that inevitably arises. Based on theoretical considerations and some simulations, it is likely that the optimal approach depends on the underlying causal structure of the data, which is typically unknown. It is also likely that the method used to assess performance may affect which approach is considered optimal. For example, recent simulations have favored very simple approaches when calibration is prioritized, but more complex approaches when discrimination is prioritized—particularly in the presence of true effect modification (72). Finally, it is uncertain whether any of these approaches will add value to the more conventional EBM approach of using an overall estimate of the main effect, or to the risk magnification approach of applying that relative estimate to a risk model.

In conclusion, we identified a large number of methodological approaches for the assessment of heterogeneity of treatment effects in RCTs developed in the past 20 years which we managed to divide into 3 broad categories. Extensive simulations along with empirical evaluations are required to assess those methods’ relative performance under different settings and to derive well-informed guidance for their implementation. This may allow these novel methods to inform clinical practice and provide decision makers with reliable individualized information on the benefits and harms of treatments. While we documented an exuberance of new methods, we do note a marked dearth of comparative studies in the literature. Future research could shed light on advantages and drawbacks of methods in terms of predictive performance in different settings.

## Data Availability

All data collected for systematic review is publically available on Medline and Cochrane Central.

## Acknowledgements

This work was supported by a Patient Centered Outcomes Research Institute (PCORI) contract, the Predictive Analytics Resource Center [SA.Tufts.PARC.OSCO.2018.01.25]. We also acknowledge support from the Innovative Medicines Initiative (IMI) and helpful comments from Victor Talisa, Data Scientist from the University of Pittsburgh.

## Notes

### Competing Interest Statement

The authors have declared no competing interest.

